# Validation of anthropometric-based weight prediction equations among Ugandan adults: A Cross-sectional study

**DOI:** 10.1101/2024.06.18.24309142

**Authors:** Zakaria Mukasa, Juliet Mutanda Ntuulo, Ronnie Kasirye, Emmanuel Olal, Christopher Lwanga, Victoria Nankabirwa, Fred Nuwaha

## Abstract

**Introduction:** Proper patient management often requires accurate weight estimation. However, the appropriate weight-measuring equipment is not always available in resource-limited settings, making clinicians resort to less reliable methods like the visual estimation of weight often with negative consequences. In this study, we assess the accuracy of anthropometric-based equations in predicting weight in Ugandan adults.

**Methods:** A cross-sectional study was conducted at Kira Health Center IV between December 2021 and February 2022. A sample of 240 adult patients, 18 years and above, was selected by quota sampling, stratified by sex and body mass index. Demographic information was obtained and anthropometric measurements including weight, height, knee height, subscapular skin fold thickness, and circumference measurements were taken. The predicted weight was computed using the proposed equations and the accuracy of the different equations was determined using Bland Altman analysis taking the equation with the best agreement as the most accurate.

**Results:** Out of 240 participants, 50% were females. The mean (standard deviation) age was 32.5(11.8) years; (34.1(12.9) years for males and 30.8(10.3) years for females). The mean (standard deviation) actual weight was 66.43 (16.33) kg while the mean height was 1.64 (0.09) meters. Using Bland-Altman analysis, Rabito equation (Weight = (0.5759xMAC) + (0.5263xAC) + (1.2452xCC) - (4.8689xS)-32.9241) was found to have the best agreement with the actual weight with a mean difference (standard deviation) of 2.55(6.99) kilograms overall.

**Conclusion:** Equation R3 was the most accurate equation for predicting weight among Ugandan adults. Therefore, in the absence of appropriate weighing scales to weigh patients, using this anthropometric-based weight estimation equation can act as a relatively accurate alternative.

## INTRODUCTION

Measurement of weight is essential for the proper management of patients (1) as safe administering of most medications requires careful tailoring of dosages by clinicians to patients’ weights to minimize overdose-related harm (2, 3). Accurate weights are essential in the effective calibration of fluid volumes, a common intervention when managing patients such as those with kidney failure, septic shock, severe dehydration, and extensive burns (4–8). Routine weight measurement in clinical care is further crucial for the diagnosis and management of conditions such as underweight, and overweight or obesity which are a growing epidemic in many countries (9).

Accurate measurement is critical to quality patient care, and yet, globally, research shows that many patients seeking care from health facilities are unweighed, hindering correct medication dosages (8, 10, 11). For example, one study in Uganda found that over 98% of patients were not weighed(11).

Failure to obtain and use accurate patients’ weights is associated with increased errors in medication dosing and subsequently suboptimal treatment, antimicrobial resistance, and drug toxicities (12–14). According to a study conducted in four hospitals in Uganda, 58% of the health workers witnessed medication errors and from a review of documents, medication errors were among the most frequent kind of errors, accounting for 17.2% of all the errors (15). In another study, drug overdose was found to be the most common medication error, accounting for 42.9% of all medication errors (16). While these studies identified medication errors, none explored the possibility of missing patient weights as a contributor to these errors. The omission of weights as a possible contributor to high medication errors in healthcare practices in Uganda sheds light on a critical oversight, which needs further exploration, for effective strategies to improve weight measurements in healthcare practices to be developed.

A review of the literature provides several barriers to routine measurement of patient weights in healthcare practices including heavy workload, inadequate staffing, and inaccurate weighing machines(10). Likewise, it is often difficult to obtain an accurate weight of bedridden patients using conventional scales. While chair or bedbound scales can be used to obtain their weight (17), these are expensive and often not available in resource-limited countries like Uganda(18). In these situations, patients’ weight is just estimated by visual estimation or using the 70 kg standard adult weight, leading to either over or underestimation.

Several weight prediction techniques have been proposed over time, for both adults and children(19–22), nevertheless, they often have low accuracy(23). Due to weight measurement challenges, anthropometric-based weight prediction has been employed for a long time both in adults(20) and pediatrics (22) and it’s more objective than visual weight estimation. Anthropometric weight predictions involve estimating a person’s weight using measurements of body size and proportions rather than directly weighing them on scales. However, weight prediction equations have mostly been developed in high-income settings(20). While there is a dearth of research in this area, a few pioneering studies have made efforts to validate and adapt weight measurement equations for the sub-Saharan African context (24, 25). One such study successfully validated the use of an anthropometric-based dose for praziquantel among children, which greatly improved access to this lifesaving medicine in rural and remote populations with high schistosomiasis endemicity (26). This tool has nevertheless a main limitation of being used only for praziquantel dosing for children. Another study tested a weight prediction equation among university students in Nigeria (27). Although their formula had good accuracy in providing weight estimates among 122 adults, it had major limitations in terms of its small sample size, limited applicability to populations with heights lower than 1.2m or greater than 2.0m, and inaccuracies in estimating underweight (27).

Accurate weight estimation methods are therefore needed for both bedbound patients and individuals in communities where access to conventional weighing scales may be limited. These methods should be easy to implement, require minimal training, and provide reliable estimates of weight for use in clinical decision-making and mass treatment campaigns. This study aimed to assess the feasibility of using anthropometric-based equations for weight estimation in resource-limited settings. To achieve this goal, we evaluated the accuracy of various equations in predicting weight among Ugandan adults.

## MATERIALS AND METHODS

### Site

To evaluate the accuracy of anthropometric-based weight estimation equations, we conducted a cross-sectional study at Kira Health Center IV, a high-volume primary healthcare facility in central Uganda, between 05 January 2022 and 21 February 2022. The center serves over 100 patients daily, with approximately half of the population being adults aged 18 years and above.

### Participants

We enrolled adult participants (18 years and older) from both inpatient and outpatient units at the health facility. Inclusion criteria included the ability to stand upright for weighing and height measurement and willingness to provide informed consent. Participants who were pregnant, had altered mental status or had amputated or immobilized limbs were excluded.

### Sample size determination

The sample size was calculated using MedCalc for Windows, version 20.215. In MedCalc, a sample size calculation for the Bland Altman plot was done. The *α* error was set at 0.05, *β* at 0.10, and the expected mean difference (MD) between methods was set at 0.8kg (28), the expected standard deviation (SD) of the differences of 1.96kg, and the maximum allowable difference between methods was set at 7kg. A sample size of 28 patients was obtained for each quota leading to a total sample size of 224 participants. Assuming a refusal rate of about (5%) and using the formula (Final sample size = Effective sample size/ (1-nonresponse rate anticipated)). The final sample size was 240 participants.

### Sampling procedure

Quota sampling was used. First, sampling was stratified by sex. Then further stratification within the strata of nutritional status of the patients using BMI (underweight, normal, overweight, and obese) was done. An equal number of participants was sampled in each stratum using convenience sampling.

### Study procedures

The health center staff identified potential participants for the different categories using BMI and these were referred to the research assistant. The research assistant approached the identified participants and told them about the study. Those who were interested in participating provided written informed consent to join the study and had their anthropometric measurements taken. Recruitment for a given stratum was stopped as soon as the target sample size (30) in the stratum was achieved. Overall recruitment was stopped when the target sample size of 240 participants was achieved.

### Variables

The dependent variable for the study was estimated weight. The independent variables were subscapular skinfold thickness (mm), height (cm), knee height (cm), calf circumference (cm), arm circumference (cm), hip circumference (cm), and abdominal circumference (cm). These were collected using a skin fold thickness caliper for subscapular skinfold thickness, a stadiometer for height, and a measuring tape for the rest.

Using an interviewer-administered questionnaire, data were also collected on other variables like; age in completed years, sex (male and female), education level (preprimary, primary, secondary, and tertiary), the reason for clinic attendance (respiratory conditions, genital-urinary, gastrointestinal, cardiovascular, and others), admission status (outpatient and inpatient), the study participants’ tribe, BMI (underweight, normal, overweight, and obese) and actual weight.

### Data Sources and Measurement

Weight (W) was measured using a well-calibrated standing weighing scale (Seca type) and the same weighing scale was used for all patients. Weight was measured with the patient standing upright and barefooted on the weighing scale placed on a firm and flat surface. The participants first removed all heavy coats, sweaters, shoes, and heavy pocket contents. The weight was measured in kilograms and to the nearest one-tenth of a kilogram (100 grams) (29).

#### Height (H) was measured with a wall mounted (Seca type) stadiometer

To ensure measurement accuracy, patients were asked to stand barefooted on a flat surface with their feet flat on the floor, heels against the wall, and the head, shoulders, and buttocks touching the wall. They were instructed to look straight ahead without lifting their chins. Just before taking the measurements, the participants were told to take a deep breath, hold it, and keep their shoulders relaxed. The headpiece of the scale was placed perpendicular to the wall and lowered until it rested on top of the patient’s head. The reading was then taken. Height was measured in centimeters and to the nearest millimeter (0.1cm) (29).

#### The Subscapular Skinfold Thickness (SST)

patients were measured standing upright facing away from the tester. They were asked to put the hand behind their backs and the inferior angle of the scapular was palpated. The tester pinched the skin at the angle, pulled it away from the underlying muscle, and using a skinfold caliper, the tester measured the skinfold thickness about one centimeter below the angle. The skinfold thickness was measured in millimeters.

#### The Abdominal Circumference (AC)

was measured between the iliac crest and the last rib through the umbilical scar(30). It was measured with the patient standing upright, in centimeters to the nearest millimeter.

#### The Calf Circumference (CC)

was measured in a standing position. The largest part of the calf was identified and used a flexible non-stretch measuring tape, not so tightly applied, to measure the circumference. The measurements were done in centimeters and to the nearest millimeter.

#### Mid-arm circumference (MAC)

also known as Mid-Upper Arm Circumference (MUAC), was measured at the midpoint of the distance between the acromion and the olecranon process on the shoulder blade and the ulna respectively, of the arm. This was done using a flexible non-stretch in centimeters and to the nearest millimeter.

#### Hip Circumference (HC)

was measured at the largest circumference around the buttocks with the patient in an upright position using a flexible non-stretch measuring tape. It was measured in centimeters and to the nearest millimeter (29).

#### Knee Height (KH)

This was measured with the patient barefooted. The knee was exposed and with the patient seated in a chair with their knee at a right angle, the assessor held the tape measure between their third and fourth fingers and placed their hand flat across the thigh of the patient at about 4cm behind the front of the knee. Then the tape measure was positioned down the leg in line with the lateral malleolus (ankle bone) to the base of the heel. The measurements were recorded to the nearest half a centimeter (31).

For uniformity, the Subscapular Skinfold Thickness, the Mid-Upper Arm Circumference, and the Calf Circumferences were taken from the right side for every patient.

### Quality Assurance and Control

A research assistant with a medical background and experience taking anthropometric measurements, was recruited. The research assistant was trained to properly use the weighing scale, stadiometer, and measuring tape to take the anthropometric measurements. The training was practical, and it lasted one week. The research assistant was also trained on the research protocol and good clinical and documentation practices. The tools were first tested and calibrated among the patients at Kira Health Center IV before beginning actual recruitment. The tools (weighing scale, stadiometer, and skinfold caliper) were also calibrated (set to zero) every morning before starting to recruit participants. Field editing of the data was done by the investigator on the day the data was collected, and correctable errors were resolved on the same day.

### Equation selection

Table 1 shows the selected equations, their sample size, study area, and the respective model coefficients of determination. Crandall and colleagues utilized only two variables in their equations(32). The rest of the equations used three or more variables and their models were more predictive. Rabito and colleagues developed a single-weight prediction model for both females and males. Although his models were highly predictive, it is imperative to validate these models for both sexes and different ages due to the morphological differences conferred by gender and age(33).

**Table 1:** Equations for weight estimation selected for evaluation in the study.

| Authors | Population | Equation | Model |
| --- | --- | --- | --- |
| Chumlea<br>(Ch) (20) | 228<br>elderly<br>(USA) | W (Female) = (MAC x 0.98) + (CC x 1.27) + (SST x 0.40) + (KH x 0.87)-62.35 | R<sup>2</sup> = 0.85 |
|  |  | W (Male) = (MAC x 1.73) + (CC x 0.98) + (SST x 0.37) + (KH x 1.16)-81.69 | R<sup>2</sup> = 0.90 |
| Rabito<br>(R) (1) | 368<br>(Brazil) | W = (0.5030xMAC) + (0.5632xAC) + (1.3180xCC) + (0.0339xSST)-43.1560 (1) | R<sup>2</sup> = 0.93 |
|  |  | W = (0.4808xMAC) + (0.5646xAC) + (1.316xCC) - 42.2450 (2) | R<sup>2</sup> = 0.93 |
|  |  | W = (0.5759xMAC) + (0.5263xAC) + (1.2452xCC) - (4.8689xS)-32.9241 (3) | R<sup>2</sup> = 0.94 |
| Crandall<br>(Cr) (32) | 1471<br>Obese<br>(USA) | W (Females) = -64.6 + (2.15xMAC) + (0.54xH) | R<sup>2</sup> = 0.55 |
|  |  | W (Males) = -93.2 + (3.29xMAC) + (0.43xH)) | R<sup>2</sup> = 0.59 |
| Lorenz<br>(L) (35) | 7000<br>general<br>population<br>(German) | W (Males) = -137.432 + (0.60035xH) + (0.785xAC) + (0.392xHC) | R<sup>2</sup> = 0.85 |
|  |  | W (Females) = -110.924 + (0.4053xH) + (0.325xAC) + (0.836xHC) | R<sup>2</sup> = 0.82 |
| MAC (cm) = Mid-arm circumference, AC (cm) = Abdominal circumference, CC (cm) = calf circumference, Subscapular Skinfold Thickness = SST (mm) and S = sex (male=1 and female=2), H (cm) = height, HC (cm) = Hip Circumference, KH (cm) = Knee Height, and W (kg) = Weight. |  |  |  |

In the study, we chose equations that contain variables that require only a measuring tape and a skinfold caliper to measure because they can easily be measured in emergency rooms, even in uncooperative patients. The equations chosen were evaluated and validated for application in the Ugandan population. The equation for estimation of weight from height developed by Kokong and colleagues in Nigeria (Estimated Weight = (Height-1)100)) was developed through observation and trial and error as stated in their study and therefore, this was not considered for evaluation(27). Equations by Donini and colleagues(34) have parameters similar to Chumlea equations so these were not evaluated as well.

### Data management

The data was collected on paper case report forms (CRFs) and then entered into Microsoft Excel. Weight predicted by each of the equations was computed using Excel before the data was exported to STATA 17 SE for further data management and analysis. For confidentiality, patient initials and unique identification numbers were used on the CRF to identify participants.

### Data analysis

Analysis was done at univariable, bivariable, and multivariable levels. Continuous variables were summarized using means and standard deviations (SD). The categorical variables were summarized using frequencies and proportions/percentages. The mean weights by the different equations were calculated and shown in Table 4 below. These predicted mean weights were compared to the actual mean weight using the paired t-test for equality of means at a 5% (*α* = 0.05) level of significance. The bias, the level of agreement (LOA), and the precision of estimates were obtained using the Bland-Altman plots with 95% Confidence Intervals (CI). These enabled the determination of the equation which was most accurate at the estimation of weight (which was the equation with the best agreement with actual weight using BA analysis). The assumptions of Bland Altman’s analysis were met before conducting the analysis. Bland Altman’s analysis is used to describe the level of agreement between two methods of the same measurements(36). It can be used to compare two new methods or to compare one method to a gold standard and it helps quantify agreement by constructing the limits of agreement using the SD and MD(37). It involves using scatter plots with the Mean of the two methods on the X-axis and the difference between the two methods on the Y-axis. Less scatter and closeness to the mean bias line represent good agreement(38).

### Ethical Considerations

Permission was sought from the Makerere University School of Public Health Higher Degrees Research and Ethics Committee. Administrative clearance was obtained from the District Health Officer (DHO) of Wakiso district and the in-charge Kira Health Center IV. Written informed consent was sought from the patients before carrying out any study procedures. Only adults were approached for consent, all participants provided consent by themselves, and none required a witness.

## RESULTS

### Description of study participants

A total of 240 patients who fulfilled the eligibility criteria were enrolled in the study. An equal number of males and females were sampled. The overall mean (SD) age of the patients was 32.48 (11.76) years: for women was 30.83 (10.28) and for men was 34.12 (12.91), with an overall age range of 18-75 years (Table 2).

**Table 2:** Summary of descriptive statistics by Sex (Continuous variables)

| Continuous Variables |  |  |  |
| --- | --- | --- | --- |
|  | All: | Females: | Males: |
|  | mean (SD) | mean (SD) | mean (SD) |
| Age (years) | 32.48 (11.76) | 30.83 (10.28) | 34.12 (12.91) |
| Weight (kg) | 66.43 (16.33) | 64.36 (17.22) | 68.5 (15.19) |
| Height (cm) | 163.7 (9.33) | 160 (8.97) | 167.3 (8.20) |
| Abdominal Circumference (cc) | 83.60 (13.72) | 84.01 (15.56) | 83.18 (11.64) |
| Mid-arm circumference (cm) | 28.61 (4.31) | 28.41 (4.83) | 28.80 (3.74) |
| Knee Height (cm) | 50.73 (4.39) | 49.09 (4.76) | 52.37 (3.27) |
| Subscapular Skinfold Thickness (mm) | 13.21 (7.16) | 14.79 (7.97) | 11.63 (5.85) |
| Calf Circumference (cm) | 35.04 (4.50) | 34.71(4.32) | 35.38 (4.66) |
| Hip Circumference (cm) | 98.46 (12.44) | 100.55 (14.29) | 96.38 (9.90) |

### Accuracy of various anthropometric-based weight estimation equations

Overall, at bivariable analysis, equation L (by Lorenz et al.) had the smallest MD of 0.72 and a p-value of 0.166. After stratification by sex, equations R2 (by Rabito et al.) and Ch (by Chumlea et al.) had the smallest MD for females and males respectively. A summary of the mean, SD, MD, and p-values is given in Table 3.

**Table 3:** Comparison between actual and estimated body weight by Sex.

|  | Mean | SD | MD | p | Mean | SD | MD | p | Mean | SD | MD | p |
| --- | --- | --- | --- | --- | --- | --- | --- | --- | --- | --- | --- | --- |
| All (Males and Females) |  |  |  |  | Females |  |  |  | Male |  |  |  |
| R1 | 64.95 | 14.64 | 1.48 | 0.002 | 64.69 | 16.04 | -0.33 | 0.518 | 65.21 | 13.15 | 3.29 | <0.001 |
| R2 | 64.82 | 14.38 | 1.61 | <0.001 | 64.52 | 15.74 | -0.16 | 0.760 | 65.13 | 12.94 | 3.37 | <0.001 |
| R3 | 63.88 | 14.22 | 2.55 | <0.001 | 61.13 | 15.30 | 3.23 | <0.001 | 66.63 | 12.51 | 1.87 | 0.010 |
| Ch | 63.03 | 13.95 | 3.40 | <0.001 | 58.19 | 13.74 | 6.17 | <0.001 | 67.87 | 12.44 | 0.63 | 0.390 |
| L | 65.70 | 14.64 | 0.72 | 0.166 | 65.30 | 15.99 | -0.94 | 0.120 | 66.11 | 13.22 | 2.39 | 0.005 |
| Cr | 78.21 | 12.36 | -11.78 | <0.001 | 82.89 | 10.47 | -18.53 | <0.001 | 73.52 | 12.38 | -5.02 | <0.001 |
| W | 66.43 | 16.33 | - | - | 64.36 | 17.22 | - | - | 68.50 | 15.19 | - | - |

**Table 4:**
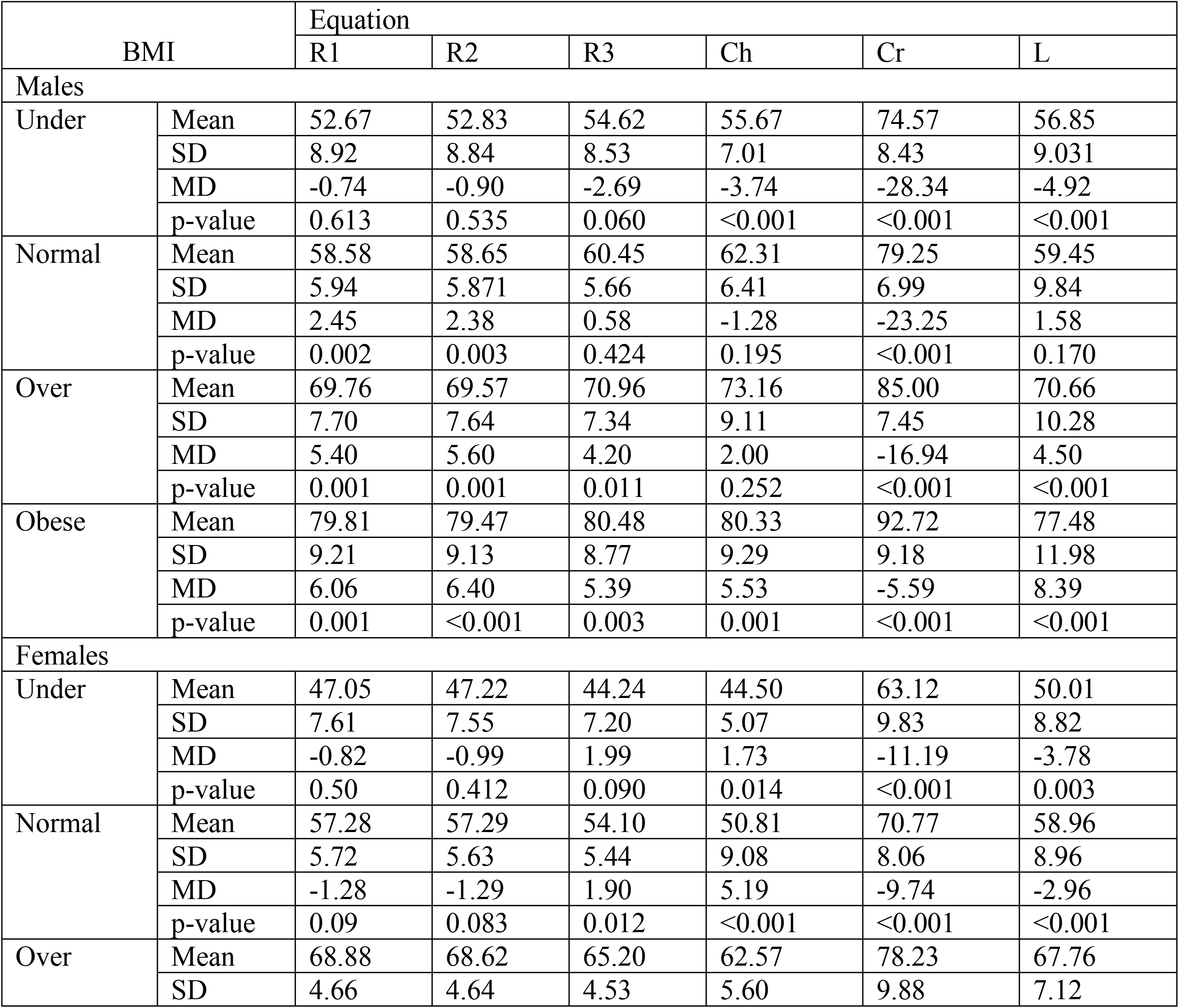
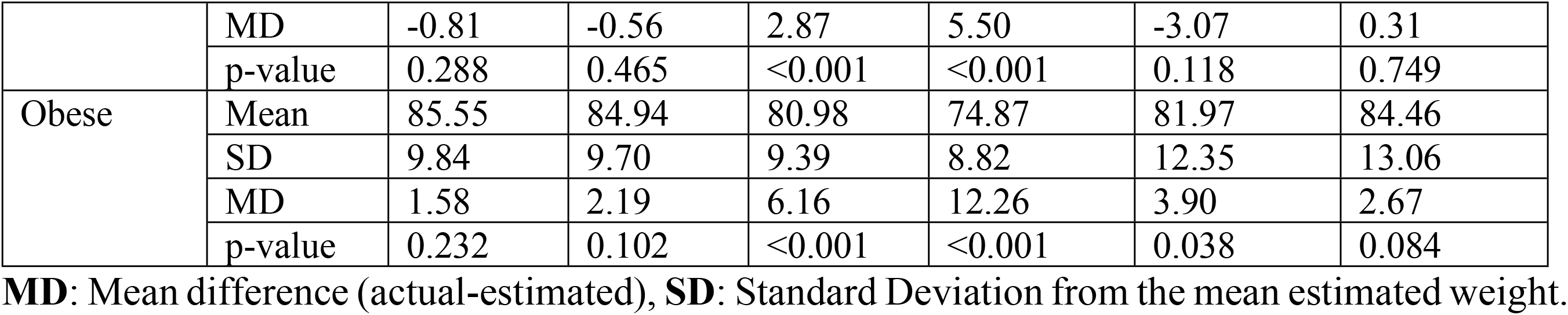
Comparison between actual and estimated body weight by BMI and Sex.

**Table 5:** Shows the coefficient of determination for the respective equations the mean difference and the standard deviation.

|  | Overall |  | Female |  | Male |  |
| --- | --- | --- | --- | --- | --- | --- |
| Equation | R-Squared | MD (SD) | R-Squared | MD (SD) | R-Squared | MD (SD) |
| R1 | 0.8061 | 1.48 (7.19) | 0.8924 | -0.33(**5.65**) | 0.7172 | 3.29(8.08) |
| R2 | 0.8046 | 1.61 (7.23) | 0.8910 | -0.16(5.71) | 0.7140 | 3.37(8.12) |
| R3 | 0.8178 | 2.55 (**6.99**) | 0.8952 | 3.23(5.66) | 0.7177 | 1.87(8.08) |
| Ch | 0.7602 | 3.40 (8.00) | 0.8482 | 6.17(7.003) | 0.7248 | 0.63(**7.98**) |
| L | 0.7560 | 0.724 (8.08) | 0.8543 | -0.94(6.57) | 0.6474 | 2.39(9.05) |
| Cr | 0.4433 | -11.78 (12.28) | 0.7134 | -18.53(10.08) | 0.5339 | -5.02(10.45) |

S1 Fig 1 shows several bland Altman plots A to F. Graph A, shows the bland Altman plot between actual weight and weight estimated by Rabito 1 (R1) equation, graph B between actual weight and estimated weight by Rabito 2 (R2), graph C between actual weight and Rabito 3 (R3), graph D between actual weight and weight estimated from Crandall (Cr) equation, graph E between actual weight and weight estimated from Chumlea (Ch) equation and graph F is for actual weight and weight estimated from equations by Lorenz et al (L). The central dashed line represents the mean difference, and the upper and lower outer parallel dashed lines represent the upper and lower limits of agreement respectively. All equations underestimated weight except the Cr equation which overestimated it. Equations R1, R2, and R3 have most of their points closer to the mean difference line as compared to the other equations. Equation Cr has the most spread-out points on the Bland Altman plots. All equations overestimated weight for patients with small weights and underestimated weight for patients with large weights.

S1 Fig 1: Bland Altman plots comparing the predicted weight by different equations and Actual weight.

Overall, equation R3 was the best at predicting weight while equation Cr was the worst. For females, equation R1 was the best at predicting weight, and equation Ch for males.

## DISCUSSION

The study evaluated the accuracy of six weight prediction equations proposed in the literature in the Ugandan population. Based on the quantitative data analysis done in the study, weight prediction from the R3 equation (by Rabito et al.) had the best agreement with the actual weight using Bland-Altman analysis. R3 is an equation that uses a simple cheap and readily available measuring tape to take the mid-arm, abdominal, and calf circumference, which can easily be measured in bedridden patients with minimal turning. When stratified by sex, equations R1 and Ch had the best agreement with actual weight for females and males respectively. However, they contained variables that require sophisticated and expensive equipment like skinfold thickness calipers and knee height calipers to measure. This would limit its applicability in resource-constrained settings. In 2008, in Brazil, Rabito and colleagues came to comparable conclusions when they evaluated anthropometric-based weight prediction equations they had developed earlier in 2006 and those that had been developed by Chumlea and colleagues(39).

In inpatient care, and more importantly for critically ill, emergency, elderly long-term bedridden patients, and in community mass drug administration, knowing the accurate body weight of a patient is important. Accurate weight is essential in critical care to minimize drug overdoses, especially for drugs with a small therapeutic window(40). Appropriate weighing scales are not always available in resource-limited settings to measure patients’ weight. Several studies have evaluated the accuracy and appropriateness of weight estimation equations, nonetheless, only a few have evaluated several equations at once like in this study where six equations were evaluated at once(28). The evaluation of several equations enabled us to have a wider number of options for selection, hence reducing bias.

In Latvia, Balode, and colleagues found the R1 equation to be the most accurate(28) which was different from the results obtained from this study, albeit using a different measure of accuracy. In this study, only equation L had a non-significant MD overall. The difference could be due to the different study populations. Their population was made of predominantly elderly females, unlike our study sample which had predominantly young adults of both sexes in equal numbers. Several studies have shown that there are gender(41, 42) and age(43) differences in anthropometric dimensions. With stratification by sex, R1 had a non-significant MD for females which is comparable to findings from Latvia. Our study used the degree of agreement measured by the Bland-Altman analysis as the measure of accuracy and found equation R3 to be the most accurate, unlike Balode and colleagues who used the mean difference. We used the Bland-Altman analysis because it provides more information about the relationship between methods than using either the paired t-test(44) or correlation(38, 45). It provides information about the magnitude and direction of the differences between the two methods of weight prediction, in addition to information about the overall agreement between the methods. In contrast, the paired t-test only provides information about the average difference between the two methods and correlation only tells us about the strength and direction of the relationship.

Generally, all equations underestimated the weight except equation Cr which overestimated the weight. This was different from what was found in Nigeria by Kokong and colleagues where their novel weight prediction method overestimated weight(27). All equations performed poorer in males than females and in obese individuals compared to those in other nutrition categories.

Overall, the data supports the theory that other anthropometric measurements can be used for predicting weight and anthropometric weight estimation equations.

### Strengths and Limitations

A major strength of this study was the stratification of study participants by nutritional status and sex. Stratification by nutritional status made it possible to obtain enough participants in the extremes of nutritional statuses that would otherwise be underrepresented with non-stratified sampling. This enabled us to assess how the proposed equations performed in these extremes. Stratifying by sex also enabled us to assess how the performance of the equations was affected by sex.

The major limitation of the study was a potential selection bias due to the use of non-probability sampling techniques. The sample ultimately obtained was predominantly young adults hence not representative of all adults and especially elderly bedridden patients on whom the equations may be applied. Stratification by sex and nutritional status was done, however, representation in the different categories of other variables like age and ethnicity was not very meaningful. Measurement error cannot be eliminated, and this may have introduced bias. However, this was reduced by using trained health workers using calibrated and standard equipment.

## CONCLUSIONS AND RECOMMENDATIONS

Equation R3 was found to have the best agreement overall and R1 and Ch for females and males respectively. In situations where patients’ weight cannot be measured, equation R3 is recommended for use to predict the weight of young adult patients with relative confidence. Equation R3 only requires a measuring tape to use which is cheap and readily available. Equations R1 and Ch require a skin fold thickness and knee height calipers to compute and therefore would not be very easily applicable in resource-limited settings. However, this equation should be used with caution in obese and underweight individuals.

Further future research should aim at evaluating these equations in the elderly population and or develop equations tailored for the Ugandan and sub-Saharan African population.

This equation could also be digitalized and incorporated into a phone-based application that can be used to predict weight.

## Data Availability

All relevant data are within the manuscript and its Supporting Information files.

## ACKNOWLEDGMENTS

We wish to acknowledge the efforts of the lecturers of Makerere University School of Public Health for their unrelenting effort and guidance throughout the course of the study. We would also like to acknowledge the assistance and support rendered to me by the DHO of Wakiso district and the administration of Kira Health Center IV. Our appreciation also goes to the patients of Kira Health Center IV for their voluntary participation that made this study a reality. We acknowledge EDCTP grant number TMA2018SF-2479, Career Strengthening for improved randomized controlled trials in Uganda, for supporting the writing process.

This study is a version of the study that was conducted for the dissertation for partial fulfillment of the requirements for the Master of Public Health from Makerere University.

## SUPPORT INFORMATION FILES

Files name: S1 Fig 1. Bland Altman plots comparing the predicted weight by different equations and Actual weight.

Files name: S2 data file.

**S1 Fig 1.**
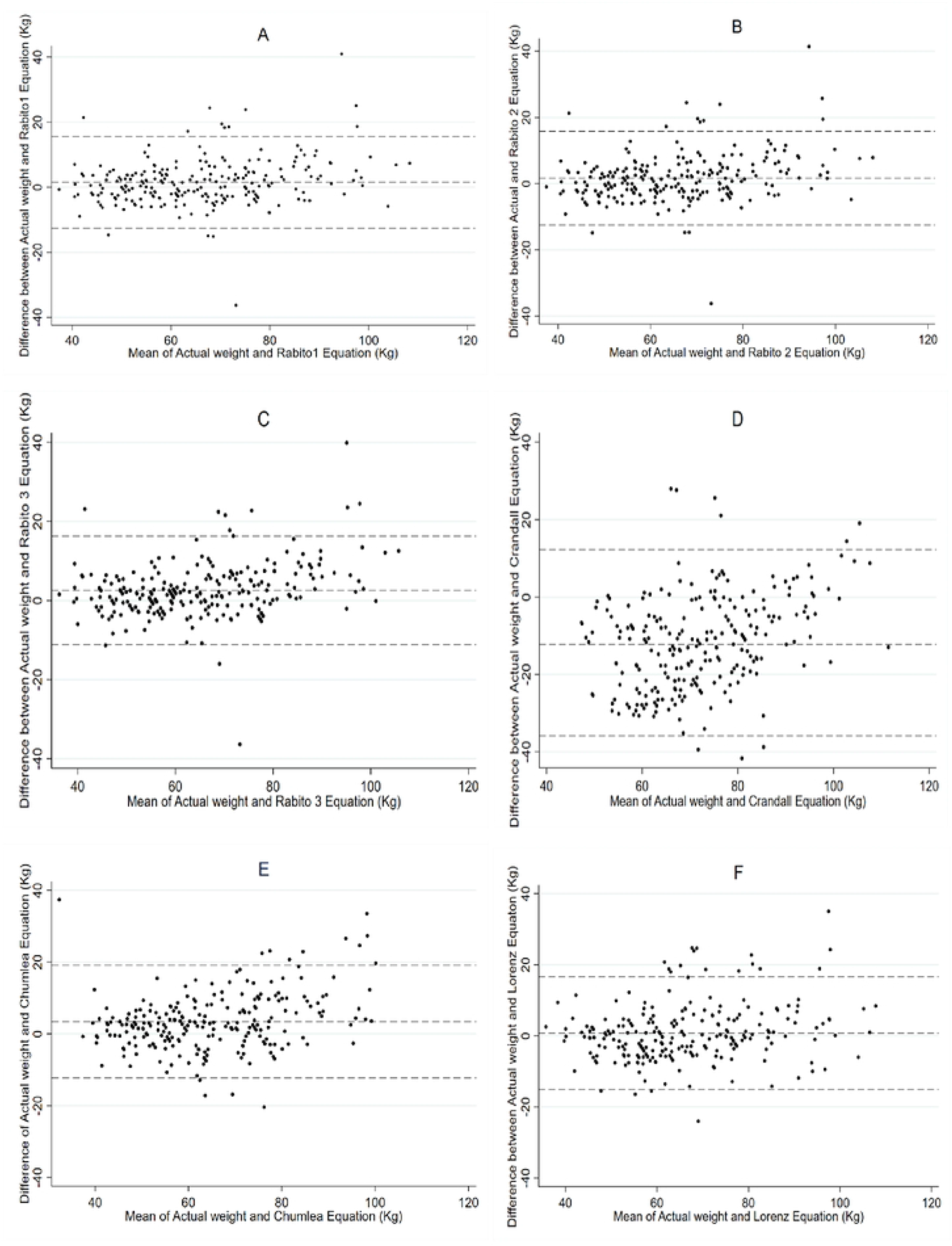
Bland Altman plots comparing the predicted weight by different equations and the Actual weight.

## Notes

### Competing Interest Statement

The authors have declared no competing interest.

### Funding Statement

Yes

### Author Declarations

Makerere University School of Public Health Higher Degrees Research and Ethics Committee

## REFERENCES

1. Rabito EI, Vannucchi GB, Suen VMM, Castilho Neto LL, Marchini JS. Weight and height prediction of immobilized patients. Revista de Nutrição. 2006;19(6):655–61.

2. Committee JF. BNF 78: September 2019-March 2020. BMJ Group/Pharmaceutical Press. 2019.

3. Blix HS, Viktil KK, Moger TA, Reikvam A. Drugs with narrow therapeutic index as indicators in the risk management of hospitalised patients. Pharmacy practice. 2010;8(1):50.

4. Toukhsati SR, Driscoll A, Hare DL. Patient self-management in chronic heart failure– establishing concordance between guidelines and practice. Cardiac failure review. 2015;1(2):128.

5. Mengesha MM, Deyessa N, Tegegne BS, Dessie Y. Treatment outcome and factors affecting time to recovery in children with severe acute malnutrition treated at outpatient therapeutic care program. Global health action. 2016;9(1):30704.

6. Committee JF. British National Formulary (online). London: BMJ Group and Pharmaceutical Press; 2015. URL: www.medicinescomplete.com (accessed 7 June 2016). 2015.

7. Goodman LS. Goodman and Gilman’s the pharmacological basis of therapeutics: McGraw-Hill New York; 1996.

8. Charani E, Gharbi M, Hickson M, Othman S, Alfituri A, Frost G, Holmes A. Lack of weight recording in patients being administered narrow therapeutic index antibiotics: a prospective cross-sectional study. BMJ open. 2015;5(4):e006092.

9. Safaei M, Sundararajan EA, Driss M, Boulila W, Shapi’i A. A systematic literature review on obesity: Understanding the causes & consequences of obesity and reviewing various machine learning approaches used to predict obesity. Computers in biology and medicine. 2021;136:104754.

10. Flentje KM, Knight CL, Stromfeldt I, Chakrabarti A, Friedman ND. Recording patient bodyweight in hospitals: are we doing well enough? Internal Medicine Journal. 2018;48(2):124–8.

11. Kavuma M, Mars M. The effect of an integrated electronic medical record system on malaria out-patient case management in a Ugandan health facility. Health Informatics Journal. 2022;28(4):14604582221137446.

12. Evans L, Best C. Accurate assessment patient weigh. Nursing times. 2014;110(12):12–4.

13. Roberts JA, Kruger P, Paterson DL, Lipman J. Antibiotic resistance—what’s dosing got to do with it? Critical care medicine. 2008;36(8):2433–40.

14. Maskin L, Attie S, Setten M, Rodriguez P, Bonelli I, Stryjewski M, Valentini R. Accuracy of weight and height estimation in an intensive care unit. Anaesthesia and intensive care. 2010;38(5):930–4.

15. Katongole SP, Anguyo RD, Nanyingi M, Nakiwala SR. Common medical errors and error reporting systems in selected Hospitals of Central Uganda. 2015.

16. Mauti G, Githae M. Medical error reporting among physicians and nurses in Uganda. African health sciences. 2019;19(4):3107–17.

17. Assured MA. Best practice: how to weigh someone who can’t stand United Kingdom: MARSDEN Accuracy Assured; [cited 2021 20 Jan 2021]. Available from: https://www.marsden-weighing.co.uk/index.php/blog/2017/10/26/best-practice-weigh-someone-cant-stand/.

18. Namale G, Kamacooko O, Makhoba A, Mugabi T, Ndagire M, Ssanyu P, et al. HIV sero-positivity and risk factors for ischaemic and haemorrhagic stroke in hospitalised patients in Uganda: A prospective-case-control study. Public Health in Practice. 2021;2:100128.

19. Cristy M. Reference man anatomical model. Oak Ridge National Lab., TN (United States); 1994.

20. Chumlea WC, Guo S, Roche AF, Steinbaugh M. Prediction of body weight for the nonambulatory elderly from anthropometry. Journal of the American Dietetic Association. 1988;88(5):564–8.

21. Anglemyer BL, Hernandez C, Brice JH, Zou B. The accuracy of visual estimation of body weight in the ED. The American journal of emergency medicine. 2004;22(7):526–9.

22. Lubitz DS, Seidel JS, Chameides L, Luten RC, Zaritsky AL, Campbell FW. A rapid method for estimating weight and resuscitation drug dosages from length in the pediatric age group. Annals of emergency medicine. 1988;17(6):576–81.

23. Kahn CA, Oman JA, Rudkin SE, Anderson CL, Sultani D. Can ED staff accurately estimate the weight of adult patients? The American journal of emergency medicine. 2007;25(3):307–12.

24. Bernal-Orozco MF, Vizmanos B, Hunot C, Flores-Castro M, Leal-Mora D, Fernández-Ballart J. Equation to estimate body weight in elderly Mexican women using anthropometric measurements. Nutrición Hospitalaria. 2010;25(4):648–55.

25. Jung M, Chan M, Chow V, Chan Y, Leung P, Leung E, et al. Estimating geriatric patient’s body weight using the knee height caliper and mid-arm circumference in Hong Kong Chinese. Asia Pacific journal of clinical nutrition. 2004;13(3):261.

26. Sousa-Figueiredo JC, Betson M, Stothard JR. Treatment of schistosomiasis in African infants and preschool-aged children: downward extension and biometric optimization of the current praziquantel dose pole. International health. 2012;4(2):95–102.

27. Kokong DD, Pam IC, Zoakah AI, Danbauchi SS, Mador ES, Mandong BM. Estimation of weight in adults from height: a novel option for a quick bedside technique. International journal of emergency medicine. 2018;11:1–9.

28. Balode A, Stolarova A, Villerusa A, Zepa D, Kalnins I, Vētra J. Estimation of body weight and stature in latvian hospitalized seniors. Papers on Anthropology. 2015;24(2):27–36.

29. Lohman TG, Roche AF, Martorell R. Anthropometric standardization reference manual: Human kinetics books; 1988.

30. Chaves TdO, Reis MS. Abdominal Circumference or Waist Circumference? International Journal of Cardiovascular Sciences. 2019;32:290–2.

31. Todorovic V, Russell C, Elia M. The ‘MUST’explanatory booklet: a guide to the ‘Malnutrition Universal Screening Tool’(‘MUST’) for adults, 2011.

32. Crandall CS, Gardner S, Braude DA. Estimation of total body weight in obese patients. Air medical journal. 2009;28(3):139–45.

33. Bredella MA. Sex differences in body composition. Sex and gender factors affecting metabolic homeostasis, diabetes and obesity. 2017:9–27.

34. Donini L, De Felice M, De Bernardini L, Ferrari G, Rosano A, De Medici M, Cannella C. Body weight estimation in the Italian elderly. JOURNAL OF NUTRITION HEALTH AND AGING. 1998;2:92–5.

35. Lorenz MW, Graf M, Henke C, Hermans M, Ziemann U, Sitzer M, Foerch C. Anthropometric approximation of body weight in unresponsive stroke patients. Journal of Neurology, Neurosurgery & Psychiatry. 2007;78(12):1331–6.

36. Bland JM, Altman D. Statistical methods for assessing agreement between two methods of clinical measurement. The lancet. 1986;327(8476):307–10.

37. Giavarina D. Understanding bland altman analysis. Biochemia medica. 2015;25(2):141–51.

38. Doğan NÖ. Bland-Altman analysis: A paradigm to understand correlation and agreement. Turkish journal of emergency medicine. 2018;18(4):139–41.

39. Rabito E, Mialich M, Martínez EZ, García R, Jordao AJ, Marchini JS. Validation of predictive equations for weight and height using a metric tape. Nutrición Hospitalaria. 2008;23(6):614–8.

40. Herout PM, Erstad BL. Medication errors involving continuously infused medications in a surgical intensive care unit. Critical care medicine. 2004;32(2):428–32.

41. Rezende FAC, Ribeiro AQ, Priore SE, Franceschinni SdCC. Anthropometric differences related to genders and age in the elderly. Nutrición Hospitalaria. 2015;32(2):757–64.

42. Silva AM, Shen W, Heo M, Gallagher D, Wang Z, Sardinha LB, Heymsfield SB. Ethnicity-related skeletal muscle differences across the lifespan. American Journal of Human Biology: The Official Journal of the Human Biology Association. 2010;22(1):76–82.

43. Su Y-J, Ho C-C, Lee P-F, Lin C-F, Hung Y-C, Chen P-C, et al., editors. Gender and Age Differences in Anthropometric Characteristics of Taiwanese Older Adults Aged 65 Years and Older. Healthcare; 2023: MDPI.

44. Van Stralen K, Dekker F, Zoccali C, Jager K. Measuring agreement, more complicated than it seems. Nephron Clinical Practice. 2012;120(3):c162–c7.

45. Ranganathan P, Pramesh C, Aggarwal R. Common pitfalls in statistical analysis: Measures of agreement. Perspectives in clinical research. 2017;8(4):187.

